# Association between asthma and memory loss: analysis from NHANES database and Mendelian randomization

**DOI:** 10.1101/2025.11.17.25340435

**Authors:** Hai Hu, Yongjun Hu, Zhonglei He, Run Huang, Xiaodi He, Wei Wei, Fangshuo Tang

**Affiliations:** First Affiliated Hospital of Anhui University of Science and Technology, Huainan, China; Joint Research Center of Vocational Medicine and Health, Great Health Research Institute, Hefei Comprehensive National Science Center,Huainan, China; Anhui University of Science and Technology, Huainan, China; Nanjing University of Information Science and Technology, Nanjing, China; Suzhou Yongding Hospital, Suzhou,China

**Keywords:** Memory loss, Asthma, Association analysis, NHANES, Mendelian randomization

## Abstract

**Background:** This study explores the link between asthma and memory decline, with a focus on Alzheimer’s disease, a disease characterized by memory loss that can be intervened with asthma medication.

**Research design and methods:** In this study, the National Health and Nutrition Examination Survey (NHANES) database combined with Mendelian randomization (MR) was utilized to explore the association between asthma and memory loss.

**Results:** The asthma was significantly associated with memory loss, both as an independent exposure factor and after adjusting for various covariates (model 1: odds ratio (OR) = 1.83, 95% confidence intervals (CI) = 1.22-2.74, p = 0.004; model 2: OR = 2.06, 95% CI = 1.34-3.16, p = 0.001; model 3: OR = 1.93, 95% CI = 1.24-3.00, p = 0.004). ROC curve indicated that asthma had a good predictive effect on memory loss (AUC = 0.717). Through MR analysis, there was a significant causal effect of asthma on memory loss (OR = 1.256, 95% CI = 1.015-1.553, p < 0.05). This was consistent with NHANES analysis results, further strengthening association between the two.

**Conclusion:** Memory loss was associated with asthma, and the risk increased with the probability of asthma. This provided valuable insights for future research.

## 1. Introduction

Memory loss, also referred to as memory deficit, denotes a decline in the ability to acquire new information and the encoding, storage, or retrieval of long-term memories in the absence of consciousness loss. It can be triggered by a multitude of factors, including advanced age, brain disorders, medication side effects, and psychological stress. Memory loss is one of the main clinical features of Alzheimer’s disease (AD), with the risk of memory loss substantially increasing with age[1]. Memory loss not only diminishes the quality of life and work efficiency but also increases the risk of dementia in the elderly[2]. Therefore, preventing and mitigating the decline of memory is of paramount importance[3].

Asthma is a prevalent chronic non-communicable disease defined by airway inflammation and hyperresponsiveness, with diverse clinical presentations and significant heterogeneity[4]. Affecting over 300 million individuals globally, asthma ranks among the most common chronic conditions in both children and adults, exhibiting noticeable variability in the types and severities of airway inflammation and remodeling[5]. Although primarily a respiratory affliction, a growing body of research suggests that asthma may be correlated with the structure and function of the central nervous system, encompassing memory and cognitive faculties[6].

Current research exploring the nexus between memory loss and asthma is relatively sparse. However, studies have indicated that montelukast, a medication utilized in asthma treatment, may exert beneficial effects in the management of AD[7]. This suggests that asthma therapeutics could potentially ameliorate memory loss, hinting at a conceivable link between the two conditions. Experimental research indicates that asthma is a risk factor for AD or dementia.[8]. However, the risk association between asthma and memory loss remains unclear. Unraveling the connection between asthma and memory loss could deepen our comprehension of the pathophysiological mechanisms underlying both conditions and pave the way for innovative therapeutic strategies in the future.

Exploring the intricate interplay between health and disease is a critical and formidable task. The National Health and Nutrition Examination Survey (NHANES)[9], designed and conducted by the National Center for Health Statistics, is a nationally representative cross-sectional survey in the United States. NHANES employs a stratified, multistage probability sampling method, providing valuable statistical data on the health and nutritional status of the non-institutionalized civilian population in the United States. The survey encompasses demographic data, anthropometric measurements, laboratory measurements, and survey questionnaire data, which are utilized not only for epidemiological and health science research but also for determining the prevalence of major diseases and the risk factors associated with these diseases[10].

In parallel, Mendelian randomization (MR) research has emerged as a novel genetic analysis method[11]. Based on the principle that genetic alleles are naturally and randomly distributed in the population, MR studies can reduce the impact of confounding factors. MR analyzes genetic variations that represent exposure factors and outcome events, thereby exploring whether there is a causal relationship between exposure factors and outcome events[12]. Compared to traditional observational studies. MR avoids some of the limitations of observational studies and randomized controlled trials in making causal inferences[13]. With the discovery of a vast number of genetic variations closely related to specific traits in biology, along with the public release of hundreds of thousands of summary data on the association between exposure, disease, and genetic variations from numerous large-scale genome-wide association studies (GWAS), these summary data enable researchers to estimate genetic associations in large sample datasets[14].

Building on this foundation, the present study aims to leverage the rich data resources of the NHANES database, combined with MR methods, to theoretically avoid residual confounding and reverse causality, further assessing the causal relationship between asthma and the risk of memory loss, and clarifying the potential impact of asthma on memory loss at the genetic level. By integrating epidemiological survey data with genetic epidemiological methods, we expect to gain a deeper understanding of the relationship between asthma and memory loss and provide a basis for developing more effective treatments and intervention measures, thereby improving patients’ quality of life and reducing the negative impact of disease on individuals and society.

## 2. Materials and methods

### 2.1 NHANES data collection

By conducting various surveys on different groups and health-related topics, the NHANES program evaluates health and nutrition of both adults and children in the United States. All participants signed written documents indicating informed consent. This analysis included 19,931 subjects from 2011 to 2014, excluding samples with missing or refused responses in the MCQ084 Additionally, relevant covariates were excluded (missing severity of diabetes, quality of sleep, or other relevant covariates in the database marked as missing, refused, or unknown). Finally, 861 subjects were recruited (Figure 1).

**Fig 1.**
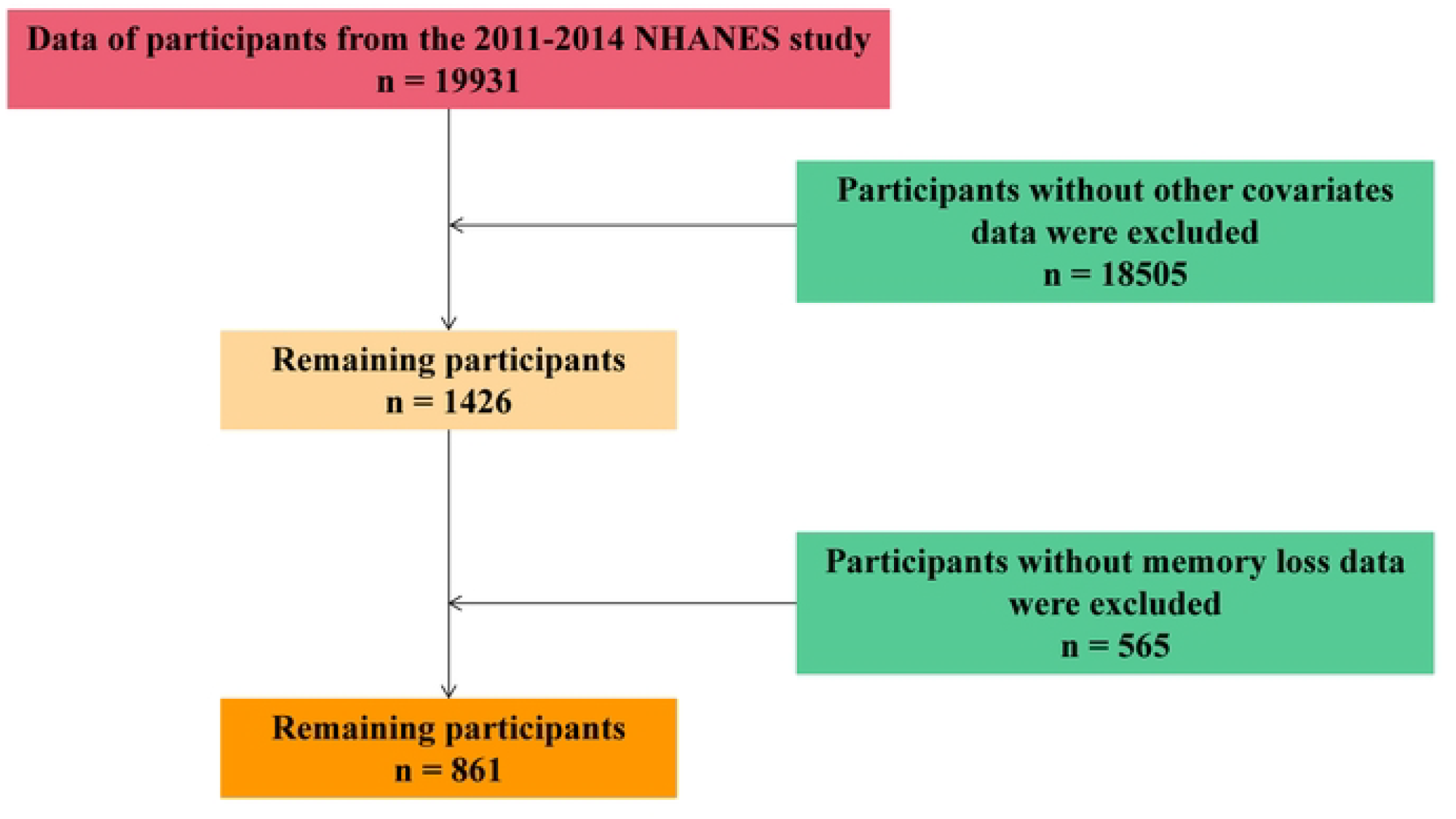

### 2.2 NHANES variables and model definition

To better evaluate NHANES data, this study defined variables and models. The outcome variable was determined by the question (MCQ084) in the questionnaire section. Those who frequently reported confusion or memory loss were classified as the memory loss group, while those who responded ‘no’ were classified as the control group.

The exposure factor was identified using the question MCQ010 from the questionnaire section. Those who were told by doctors or other healthcare professionals that they had asthma were categorized as the disease group with asthma, while those who answered ‘no’ were categorized as the control group without asthma.

To assess the impact of potential confounding factors, several important covariates were selected: race; age; gender; education level; severity of diabetes: subjects were divided into severe diabetes (yes) and regular diabetes (no) groups based on whether the doctor informed them that diabetes affected their eyes or if they had retinopathy; quality of sleep: subjects were divided into the sleep difficulty group (yes) and the non-sleep difficulty group (no) based on whether they informed a doctor or other health professional about having sleep difficulties.

Based on the confounders, the NHANES disease models were defined as follows: model 1 (no adjustment): the relationship between asthma and memory loss; model 2 (minimally adjusted model): adjusted for gender, education level, race, and age (based on model 1); model 3 (fully adjusted model): further adjusted for severity of diabetes, and quality of sleep, in addition to model 2. Variables were summarized (Table 1).

### 2.3 NHANES data analysis

In this study, baseline characteristics were described with categorical variables presented as percentages. Weighted chi-square test was applied to analyze categorical variables. To further investigate the impact of covariates on this association, relationship between asthma and memory loss was explored by calculating adjusted odds ratios (OR) and 95% confidence intervals (CI) through a weighted multivariable logistic regression model. Additionally, weighted logistic regression was employed to perform risk stratification analysis on several covariates, in order to assess the adjustment effect. The pROC package(v 1.18.5)[15] was used to plot receiver operating characteristic (ROC) curve for model 3 to evaluate predictive effect of asthma on risk of memory loss. We then utilized nhanesA package (v 1.0.3)[16] for all NHANES data analysis, with p < 0.05 considered statistically significant.

### 2.4 MR data processing and analysis

Subsequently, we performed a MR analysis to investigate causal link between asthma and memory loss. The asthma GWAS dataset finn-b-ASTHMA_MODE was sourced from IEU OpenGWAS database, which included 16,380,170 Single Nucleotide Polymorphisms (SNPs) from 155,386 European samples (asthma: control = 19,573: 135,813). In IEU OpenGWAS database, we also obtained the finn-b-MEMLOSS for memory loss outcome variable, which included 16,380,454 SNPs from 211,717 European samples (memory loss: control = 1,384: 210,333).

The R package TwoSampleMR (v 0.6.0)[17] was utilized to read exposure factors and identify SNPs that were independently associated with these factors for obtaining SNPs. These SNPs were subsequently utilized as instrumental variables (IVs). The criteria for SNP selection were as follows: a. p < 5×10−8; b. SNPs significantly associated with exposure factors were identified, using clump=TRUE, r2=0.001, kb=10000, to remove instrumental variables (SNPs) with linkage disequilibrium (LD); c. The SNPs of exposure factors and outcomes were aligned through harmonise_data function (excluding SNPs with F-statistics < 10).

We next harmonized effect alleles and effect sizes for obtaining exposure factors. MR analysis was then performed using 5 algorithms: inverse variance weighted (IVW)[18], weighted median[19], MR egger[20], weighted mode[21], and simple mode[17]. Subsequent analysis results were based on the IVW method (p < 0.05). To assess the correlation between exposure factors and outcome, scatter plots were drawn combining SNP-exposure effects and SNP-outcome effects. Forest plots were generated for assessing diagnostic efficiency of each SNP locus for outcome. Funnel plots were drawn to determine the randomness of the analysis and whether MR adhered to Mendel’s second law of random assortment, combining standard error (SE) and β of each IV. Sensitivity analysis was conducted in order to assess the reliability of MR results. The mr_heterogeneity function was first applied for heterogeneity test, with a threshold of p > 0.05 indicating no heterogeneity. In this study, pleiotropy test was tested using the mr_pleiotropy_test function from TwoSampleMR package (v 0.6.0) and the mr_presso function from MRPRESSO package (v 1.0)[22], with a threshold of p > 0.05 indicating no horizontal pleiotropy. In addition, mr_leaveoneout function from the TwoSampleMR package (v 0.6.0) was utilized to conduct leave-one-out (LOO) analysis to observe whether each SNP significantly altered the outcome. Finally, to verify that the results of the forward analysis were not affected by reverse causality, the directionality_test function was utilized to perform MR Steiger directionality analysi, with asthma as exposure factor and memory loss as outcome.

### 2.5 Statistical analysis

The R programming language (v 4.2.2) was used to conducted bioinformatics analyses. We compared the relationships between groups and the mean differences using the weighted chi-square test and the T-test, respectively, with p < 0.05 considered statistically significant.

## 3. Results

### 3.1 Description of baseline characteristics of study subjects

The results showed that after stratifying the outcome by the presence of memory loss (No for not having memory loss, Yes for having memory loss), among all samples, 42 individuals had both asthma and memory loss, 98 had asthma but not memory loss, 137 had memory loss but not asthma, and 584 had neither disease. Asthma exhibited a significant effect on memory loss (p < 0.01). In addition, all covariates showed significant intergroup differences (p < 0.05), suggesting that all covariates also exhibited a significant effect on the memory loss (Table 2).

### 3.2 The association between asthma and memory loss using NHANES data analysis

To investigate whether the OR between asthma and memory loss changed significantly with the addition of covariates, two adjusted models were constructed, assuming that all covariates interacted with memory loss. Then, three consecutive multivariate generalized linear model (GLM) regression models were constructed. The results indicated that the OR values were consistently greater than 1 in all models and p values were consistently less than 0.05 (model 1: OR = 1.83, 95% CI = 1.22-2.74, p = 0.004; model 2: OR = 2.06, 95% CI = 1.34-3.16, p = 0.001; model 3: OR = 1.93, 95% CI = 1.24-3.00, p = 0.004), suggesting that the effect of memory loss on asthma was not significantly confounded by other covariates, and that asthma was a risk factor for memory loss (Table 3).

To further confirm the stability of the correlation between asthma and memory loss risk in different populations, a risk stratification analysis was conducted (based on model 3). The findings indicated that asthma and memory loss remained strongly correlated, with asthma being a risk factor for memory loss (OR = 1.93; 95% CI: 1.24-3.00; p = 0.004).(Figure 2A). Subsequently, ROC curve showed that asthma had a good predictive effect on memory loss (area under the curve (AUC) = 0.717). (Figure 2B).

**Fig 2.**
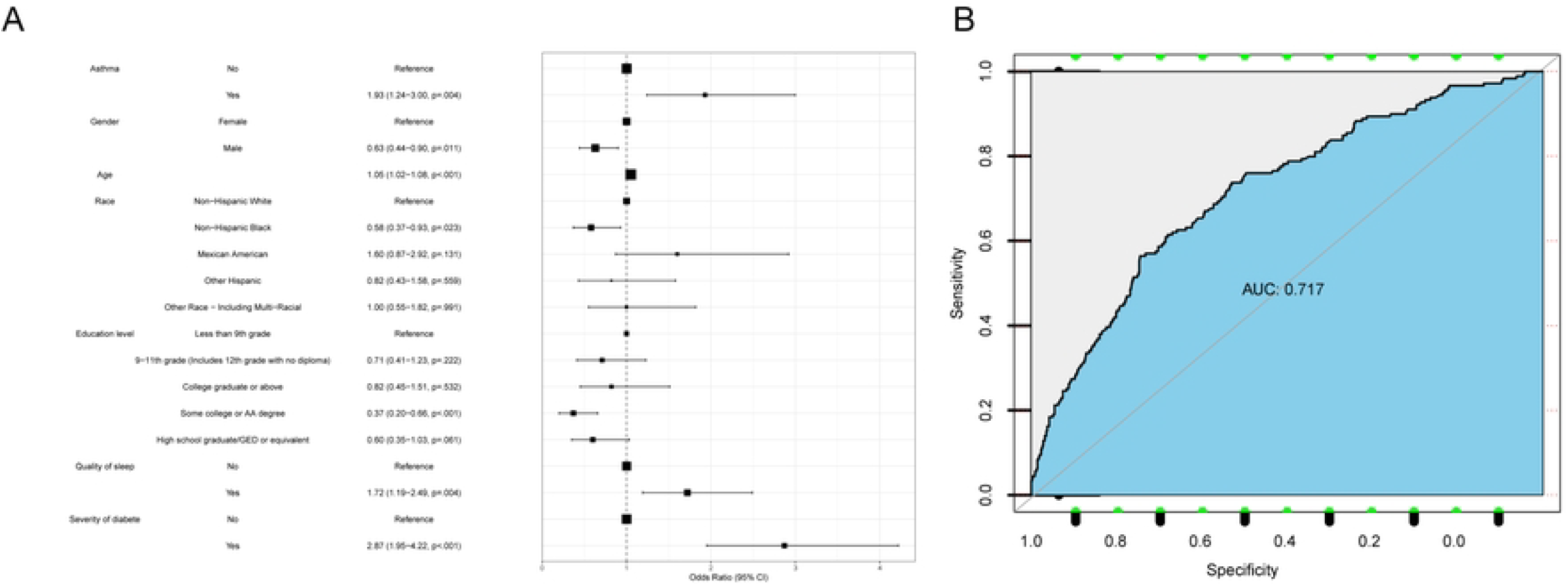

### 3.3 The unidirectional causal relationship between asthma and memory loss revealed by MR analysis

Through MR analysis, the results indicated that asthma was a risk factor and a significant causal relationship between asthma and memory loss was observed (OR = 1.255, 95%CI = 1.015-1.553, p < 0.05, based on IVW) (Table 4). Next, the scatter plot results indicated that the intercept value of the fitted line was close to 0 (based on IVW algorithm), suggesting that the correlation between the exposure factor and the outcome was minimally influenced by confounding factors, thereby maintaining the reliability of the results. Additionally, since the slope of the fitted line was positive, it indicated that asthma was a risk factor for memory loss (Figure 3A). Subsequently, the forest plot results indicated that the IVW effect size was positioned to the right of the reference line (all IVW > 0, all MR egger > 0), suggesting that asthma was a risk factor for memory loss (Figure 3B). The funnel plot results of the randomness indicated that the samples were approximately symmetrically distributed along both sides of the reference line (IVW), which was consistent with Mendel’s second law of independent assortment (Figure 3C). Subsequent sensitivity analysis indicated no heterogeneity among the samples (p > 0.05) (Table 5). In addition, to assess the presence of confounding factors, horizontal pleiotropy was tested, and the results indicated no horizontal pleiotropy and thus no confounding factors (p > 0.05) (Supplementary Table 1). At the same time, effect of the remaining SNPs on memory loss variable was minimal by sequentially excluding each SNP, indicating that the analysis results were reliable and stable (Figure 3D). Finally, MR Steiger directionality test showed that snp r2.exposure was greater than snp r2.outcome, and the correct causal direction was TRUE, with a p-value < 0.0001, suggesting a unidirectional causal relationship between asthma and memory loss (Supplementary Table 2).

**Fig 3.**
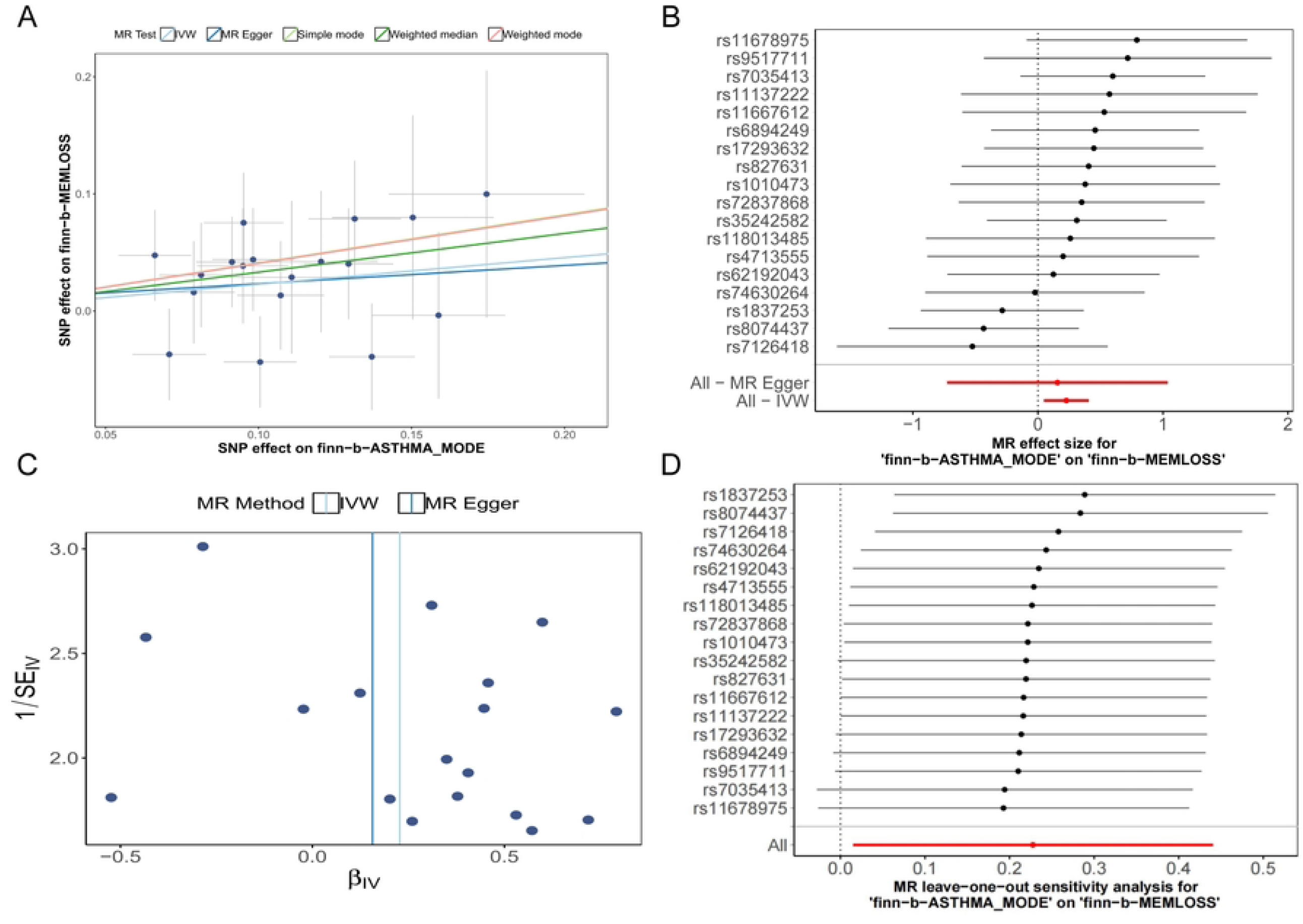

## 4. Discussion

Memory loss, as the condition progresses, patients gradually lose other cognitive functions, such as language, calculation, and judgment abilities, ultimately affecting their daily living capabilities. The pathogenesis of cognitive decline may involve various factors, including genetics, environment, lifestyle, and pathological changes in the brain. Understanding these mechanisms is crucial for developing prevention and treatment strategies[23-25]. In the research process, data from NHANES outlined baseline characteristics, association analyses, and risk stratification analyses results all suggested that asthma was significantly associated with memory loss, with the ROC curve showing a good predictive effect for asthma on memory loss. MR analysis also confirmed a positive causal relationship between asthma and memory loss.

When analyzing the confounding factors associated with the link between asthma and memory loss, we considered diabetes and sleep quality, both of which have been reported to be correlated with memory loss[26,27] and are also more common in asthma patients. Diabetes can lead to microvascular and macrovascular complications, affecting cerebral blood flow and subsequently cognitive function. Poor sleep quality directly affects the brain’s recovery and memory consolidation processes. By controlling these variables, we aim to more accurately assess the direct association between asthma and memory loss.

In exploring the potential association between memory loss and asthma, we initially reviewed previous studies, which reported animal experiments demonstrating the impact of chronic asthma on cognitive dysfunction[28], subsequently confirmed in human clinical studies that individuals with poorly controlled asthma may experience a decline in cognitive function, particularly in memory and attention[29]. During acute exacerbations or severe asthma attacks, oxygen levels can drop, leading to diffuse cerebral hypoxia, hypoxic brain damage, and changes in baseline arterial oxygen saturation, thereby affecting cognitive function[30,31]. Furthermore, studies have found that in children with asthma and obesity, serum magnesium levels are significantly reduced, and magnesium is a key mineral for maintaining normal neuronal function; low magnesium levels are associated with the loss of memory and attention[32]. Montelukast, a leukotriene receptor antagonist widely used to treat asthma, has been shown in recent studies to potentially have therapeutic effects on AD[33], improving cognitive function in AD patients[34]. Montelukast improves cognitive function by inhibiting leukotriene receptors, reducing inflammatory responses[35]. Airway hyperresponsiveness (AHR) and airway inflammation are the primary pathophysiological characteristics of asthma, with airway inflammation and/or airway remodeling potentially being key determinants of the direct stimulus response[36]. Systemic inflammation has been associated with an increased risk of dementia, including a significant component of AD pathology, as early as the early stages[37]. In addition, microglia, by memorizing peripheral inflammation and leading to adverse pathological exacerbation years later, are missing[38]. This suggests that asthma treatment drugs may have a positive impact on cognitive function, and there may be a common inflammatory mechanism between asthma and memory loss.

Furthermore, we hypothesize that there may be common molecular pathways between asthma and memory loss. For instance, chronic inflammation caused by asthma may lead to a decrease in serum magnesium levels, and magnesium deficiency can affect the excitability and synaptic plasticity of nerve cells, which are the foundations of learning and memory formation[39]. Nuclear factor kappa-B (NF-κB) produced during the inflammatory process of asthma has been shown to affect cognitive function[8]. The asthma treatment drugs Montelukast, Myrtenol, and ibudilast can improve cognitive impairment by reducing the expression of NF-κB[35,40,41], suggesting it may be a shared pathological pathway for both diseases.

Asthma has been indicated as not only a highly associated factor for memory decline but also as having a causal effect, according to combined NHANES and MR analysis. However,this study has several limitations. First, the cross-sectional nature of the study limits our ability to assess the temporal progression of these findings; second, although the NHANES data are representative, they are still based on a U.S. sample. Therefore, the results may not be applicable to populations from other countries or cultural backgrounds. Third, the NHANES data may not be updated in real-time, which could affect the timeliness of the study results. Lastly, although MR analysis can reduce the impact of confounding factors, it cannot completely exclude all possible confounders, especially those that are unmeasured or unidentified.

In summary, the relationship between asthma and memory loss is likely to be multifaceted, involving inflammation, changes in nutrient factor levels, and neurobiological alterations. Future research needs to further explore these potential mechanisms and assess the potential benefits of asthma treatment in improving cognitive function. This study provides evidence of a possible association between asthma and declining memory function. These findings provide new directions for future research and may help develop new treatment strategies to improve the overall health and cognitive function of asthma patients.

## Data Availability

All relevant data are within the manuscript and its Supporting Information files.

## Supporting information

**S1 Fig. Schematic diagram of subject selection for NHANES survey from 2011 to 2014**.

**S2 Fig. Model 3-Based Risk Stratification Analysis. (A) Risk stratification analysis chart (based on Model 3). (B) ROC curve based on Model 3**.

**S3 Fig. Comprehensive Diagnostic Model Evaluation. (A) Correlation analysis scatter plot**.. **(B) Forest plot of diagnostic efficacy. (C) Funnel plot (D) Leave-one-out sensitivity test**.

**S1 Table. Variable and its numbering information**

**S2 Table. Characteristics of NHANES participants**

**S3 Table. Correlation analysis of participants**

**S4 Table. Results of MR randomized analysis of different methods S5 Table. MR heterogeneity test results**

**Supplementary Table1. MR horizontal pleiotropy test results**

**Supplementary Table2. MR Steiger directionality test**

